# Can people with asymptomatic or pre-symptomatic COVID-19 infect others: a systematic review of primary data

**DOI:** 10.1101/2020.04.08.20054023

**Authors:** Nelson Aguirre-Duarte

## Abstract

Asymptomatic but infectious people have been reported in many infectious diseases. Asymptomatic and pre-symptomatic carriers would be a hidden reservoir of COVID-19.

**Aim:** This review identifies primary empirical evidence about the ability of asymptomatic carriers to infect others with COVID-19 pandemic and reflects on the implications for control measures.

**Methods:** A systematic review is followed by a narrative report and commentary inclusion criteria were: studies reporting primary data on asymptomatic or pre-symptomatic patients, who were considered to have passed on COVID-19 infection; and published in indexed journals or in peer review between January 1 and March 31, 2020.

**Results:** Nine articles reported on 83 asymptomatic or pre-symptomatic persons.

**Conclusions:** The evidence confirms COVID-19 transmission from people who were asymptomatic at the time. A series of implications for health service response are laid out.

## Introduction

Asymptomatic but infectious people have been reported in many infectious diseases (Fraser, Riley, Anderson, & Ferguson, 2004; Jartti, Jartti, Peltola, Waris, & Ruuskanen, 2008). Examples include herpes, cytomegalovirus and respiratory diseases caused by viruses such as H1N1 or H5N1 influenza (Fielding, Kelly, & Glass, 2015; Gu et al., 2011; Khanna, Kumar, Gupta, & Kumar, 2012; Le et al., 2013; Powell et al., 2012). Asymptomatic and pre-symptomatic people are a reservoir hidden for detection systems.

Coronavirus disease 2019 (COVID-19), has infected more than 857 thousand people globally, resulting in more than 42 thousand deaths in a period of three months according to John Hopkins website (Dong, Du, & Gardner, 2020).

The Chinese government first reported the infection as pneumonia of unknown origin in late December 2019; then in late January 2020, the WHO declared the outbreak a Public Emergency of International Concern, and on 11 March was elevated the alert level to pandemic (WHO, 2020a, 2020b). The epicentre of the disease was Wuhan City in Hubei Province in central China. Three characteristics were identified in the infection: an 83% attack rate declared as highly alarming, a wide spectrum of clinical manifestations in the patients (ranging from minor symptoms to pneumonia and death) along with evidence of the virus in a minority without any evidence of symptoms. The infection quickly spread in China and then cases began to be exported internationally (Mizumoto, Kagaya, & Chowell, 2020; Wu et al., 2020).

There is increasing evidence supporting the contribution to the spread of infection by asymptomatic and pre-symptomatic patients with COVID-19. The speed with which the infection spreads suggests that transmission from unidentified (because asymptomatic) cases plays a substantial role in the exponential increase in the number of infected patients. This review seeks empirical evidence about the presence of asymptomatic and pre-symptomatic carriers in the current COVID-19 pandemic and offers reflections on the implications in the context of the current measures taken to control it.

## Method

This study reports a systematic literature review, a tabular summary of primary studies, and a narrative commentary on the implications. Databases searched were: MEDLINE, PubMed and Google academic (which includes pre-peer reviewed articles) using search terms “coronavirus”, “COVID-19”, “asymptomatic patients”, and “pre-symptomatic patients”. Inclusion criteria were reports of primary data pointing to infection from contact with asymptomatic or pre-symptomatic persons; publication in indexed journals or in the process of peer review between January 1 and March 31, 2020. The references of included articles were scanned for further relevant sources (snowball process). Publications in English, Spanish, Mandarin and Korean were accepted.

References were managed using EndNote v8 and MAXQDA v11(Oliveira, Bitencourt, Teixeira, & Santos, 2013).

## Results

Two hundred eight abstracts were identified as potentially relevant, reduced to nine finally included, as detailed in Figure 1. reviewed, and the selection criteria applied. Twenty-five scientific articles were selected for content analysis.

**Fig.1.**
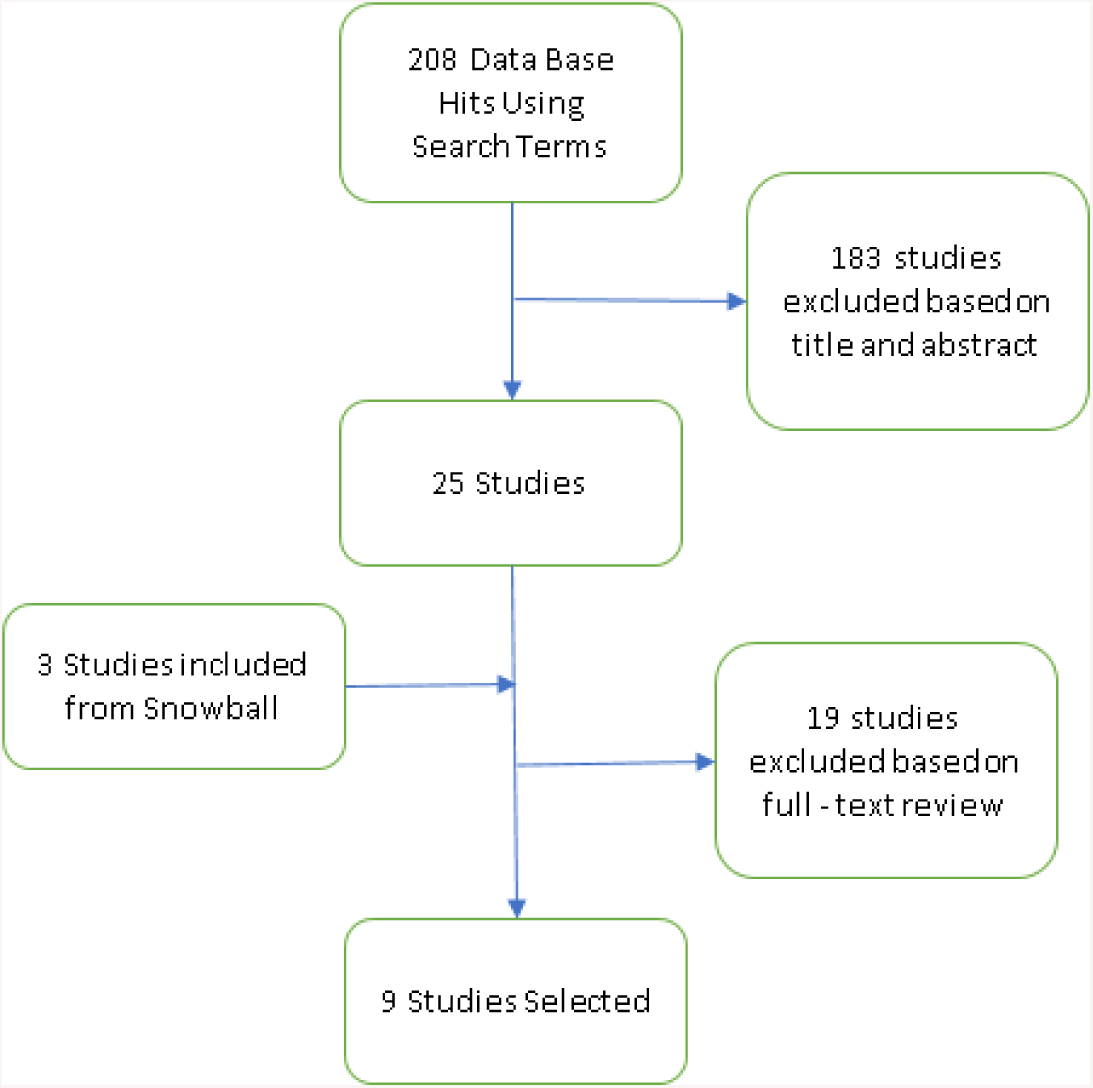
Description of the study selection process

Through a review of bibliographic references, seven new articles were included (Fig 1). A total of nine articles were identified as empirical evidence of the presence of asymptomatic or pre-symptomatic patients with COVID-19 infection. Table 1 presents the current evidence.

**Table 1.** Current evidence of asymptomatic and pre-symptomatic COVID-19 patients

| Date / period of the presentation / exposure | N asymptomatic or pre-symptomatic | Age (years) | Gender | Residence country/city of asymptomatic patient | Place of diagnostic | Context | Type of patient | Reference |
| --- | --- | --- | --- | --- | --- | --- | --- | --- |
| Dec 8, 2019 – February 6 2020 | 1 | 0.7 | Female | Wuhan/China | Wuhan/China | Retrospective study on all hospitalised infants diagnosed with COVID-19. 1 patient was asymptomatic over 9 patients. This patient is related to 5 infected family members. | Asymptomatic | (Wei et al., 2020) |
| Jan 10, 2020 | 1 | 20 | Female | Wuhan/China | Anyang/China | This woman from Wuhan visited her family in Anyang. Then her family presented symptoms and x were confirmed infected. The woman was positive for COVID-19 PCR, but she never presented symptoms despite having had multifocal ground-glass opacities on chest CT, with subsegmental areas of consolidation and fibrosis. | Asymptomatic | (Bai et al., 2020) |
| Jan 10, 2020 | 1 | 10 | Male | Shenzhen/China | Wuhan/China | This study reports clinical results of a family cluster of 5 people infected with COVID-19. The patient referred as 5, was asymptomatic despite ground glass opacities identified by CT scan. | Asymptomatic | (Chan et al., 2020) |
| Jan 11 to Feb 29, 2020 | 55 | Ages 2 to 69 years, median 49 years. The proportion of asymptomatic cases aged from 30 to 49 years old was 30.9% | 22 Male<br>33 F | Shenzhen/China | Shenzhen/China | This a retrospective analysis on asymptomatic patients admitted to a hospital after their family member's diagnoses of COVID-19. All patients were asymptomatic at the time of the test. 14 mild, 39 moderate and 2 severe during hospitalization | Pre-symptomatic | (Y. Wang et al., 2020) |
| Jan 19, 2020 | 1 | NI | Female | Shanghai | Germany | This asymptomatic woman attended business meetings in Germany. Then 4 people were COVID-19 PCR positive. | Pre-symptomatic | (Rothe et al., 2020) |
| Jan 28 to Feb 9, 2020 | 24 | Ages 5 to 95 years old. (median 32.5) | 8 Males<br>16 F | Nanjing/China | 33.3% had history of recent travel to Hubei | Investigation of all close contacts of COVID-19 patients, both in clinic and community in one city. All 24 cases were identified as asymptomatic carriers. | Asymptomatic | (Hu et al., 2020) |
| Jan 29, 2020 | 5 | NI | NI | Japan | Wuhan/China | 565 Japanese citizens were evacuated from Wuhan. Healthy individuals were in quarantine. All people were screened for symptoms prior to disembarkation. All tested for COVID-19 PCR. 5 of them were positive and asymptomatic at the time of the test. | Asymptomatic | (Nishiura et al., 2020) |
| Feb 5, 2020 | 255 | (0-9)1, (10-) 1, (20-) 2, (30-) 5, (40-)7, (50-)22, (60-)56, (70-)136, (80-)25, (90-)0 | NI | Multiple precedence – diamond Princess Cruise Ship | Yokohama/Japan | 521 people tested positive for COVID-19 over 3711 passengers and crew members. “the crude asymptomatic ratio, a simple proportion of asymptomatic infections among all the infections was estimated as follows: 66.7% (95% Confidence Interval (95%CI): 9.4%, 99.1%) for aged 0-19 years, 30.8% (95%CI: 22.6%, 40.0%) for aged 20-58 years and 52.8% (95%CI: 47.9%, 57.5%) for aged 60 years and older (95%CI is based on binomial distribution).” | Asymptomatic | (Mizumoto & Chowell, 2020) |
| Feb 6, 2020 | 1 | 50 | Female | Anqing/China | Anqing/China | This woman had a test for COVID-19 as her husband's close contact with positive patients. She had a confirmed positive result. She also received treatment with antivirals and five days after treatment both throat and anal swaps were still positive. This patient was an asymptomatic positive carrier. | Asymptomatic | (Luo et al.)* |

NI = No Informed
\*Study publish in ChinaXiv.org

## Discussion

This review presents the available evidence that asymptomatic and pre-symptomatic people can infect others with COVID-19. As for other viral infections, this may play an essential role in community transmission. The specific of proactive interventions should be considered during a pandemic that includes asymptomatic transmission.

### Implications for risk management and preparedness

it seems that the lessons learnt from previous pandemics have not been applied, and countries did not adequately assess the risks, and as a result, there has been a lack of resources and response plans adequate to save lives (Anderson, Heesterbeek, Klinkenberg, & Hollingsworth, 2020). The presence of asymptomatic and pre-symptomatic carriers imposed an extra challenge on how to protect the public in general and front-line workers from the infection when an undetected infective person is present. A shortage of medical supplies is not uncommon in public health emergencies. Therefore, national and international medical supplies programmes before emergencies arise need to be strengthened (Wong, Leo, & Tan, 2020).

### Implications for healthcare workers protection and health systems response

Asymptomatic and pre-symptomatic patients are a source of active risk for front line workers. It imposes an additional challenge to the health system as undetected patients can spread the infection and affect the level of response (J. Wang, Zhou, & Liu, 2020). The use of personal protective equipment by all front-line workers should be mandatory. Shortages of masks, goggles are unacceptable because the risk of infection and death of these workers may affect the whole health system response to the community (Schwartz, King, & Yen, 2020).

### Implication for infection modelling and planning

Mathematical models in the context of COVID-19 have demonstrated their usefulness in simulating transmission trends, risk assessment, epidemic development, and the effects of isolation and quarantine (Tang, Xiao, Peng, & Shen, 2020). However, missing the effect of asymptomatic or pre-symptomatic stages of the disease, introduces errors into prediction models (Gaeta, 2020; Tian et al., 2020).

### Implications for testing approaches

A rapid, systematic and widespread approach to testing is needed to detect new cases in the community and in front-line workers. Maintaining restrictions in testing is not the best way to identify asymptomatic and pre-symptomatic patients (C. J. Wang, Ng, & Brook, 2020). The worst-case scenario is to have asymptomatic or pre-symptomatic COVID-19 front-line workers because they can potentially infect vulnerable patients and their families. The testing approaches must be easily adaptable for the needs of the community and clusters of interest. The speed of the detection and fast response could be a critical success factor (MacIntyre & Heslop, 2020) (Ferretti et al., 2020)

### Implications for immigration restrictions

Temperature scanners and self-report of symptoms at international airports must necessarily fail to detect asymptomatic and pre-symptomatic patients. Self-isolation plus more systematic testing could prevent undetected dissemination. A more robust public health response must be developed in the border.

Special attention needs to be paid to front-line workers in airlines and other crew members. Asymptomatic patients could be a source of infection to these people and airline crew who themselves asymptomatic carriers are a risk to passengers. Airline companies should reflect on how to protect the crew and minimise the risk of dissemination (Mouchtouri, Dirksen-Fischer, & Hadjichristodoulou, 2020), probably by routine testing and use of personal protective equipment.

### Implications in social connectedness (physical distance and clusters)

Physical distance is a paramount approach to avoid COVID-19 infection, but the presence of asymptomatic patients in the community and front-line workers should be enough reason to wear face mask and goggles in public and healthcare places. The virus is present in saliva, which could be spread in the form of exhaled air (over a short distance) or droplets (over a longer distance (Bourouiba, 2020). At a community level, transmission undetected infective persons are highly likely.

These strategies should apply until a specific vaccine is available, even if effective treatments become available first. This pandemic has changed the way we interact as individuals and how to respond as communities to a severe challenge to public health. The number of deaths indicates there is substantial room for improvement is significant in the way governments and global health organisations need to operate. Next time, we would aim to more rapidly identify and disseminate information about a cluster of new disease, and a more coordinated and faster global response could isolate such a cluster and prevent rapid dissemination to the entire world.

## Limitations

Those studies selected in this review had several limitations. Many have a small sample size. Several studies note the low sensitivity or specificity of available diagnostic tools; relevant symptoms were difficult to identify in some patients (for the infant study case in particular); and in some cases, relevant dates were while there were reporting delays with others. However, given the recent presentation of this disease and the lack of formal studies in this short period, this information is relevant to public health policy and protection for populations and healthcare and other front-line workers.

## Data Availability

All references are available by request.

## Acknowledgement

I wish to thank Dr T Kenealy for a helpful review of this manuscript.

